# Stability of plasma metabolomics over 10 years among women

**DOI:** 10.1101/2022.01.05.22268819

**Authors:** Oana A. Zeleznik, Clemens Wittenbecher, Amy Deik, Sarah Jeanfavre, Julian Avila-Pacheco, Bernard Rosner, Kathryn M. Rexrode, Clary B. Clish, Frank B. Hu, A. Heather Eliassen

## Abstract

**Background:** In epidemiological studies, samples are often collected long before disease onset or outcome assessment. Understanding the long-term stability of biomarkers measured in these samples is crucial. We estimated within-person stability over 10 years of metabolites and metabolite features (N=5938) in the Nurses’ Health Study (NHS): The primary dataset included 1880 women with 1184 repeated samples donated 10 years apart while the secondary dataset included 1456 women with 488 repeated samples donated 10 years apart.

**Methods:** We quantified plasma metabolomics using two liquid chromatography mass spectrometry platforms (lipids and polar metabolites) at the Broad Institute (Cambridge, MA). Intra-class correlations were used to estimate long-term stability (10 years) of metabolites and were calculated as the proportion of the total variability (within-person + between-person) attributable to between-person variability. Within-person variability was estimated among participants who donated two blood samples approximately 10 years apart while between-person variability was estimated among all participants.

**Results:** In the primary dataset, the median ICC was 0.43 (1^st^ quartile [Q1]: 0.36; 3^rd^ quartile [Q3]: 0.50) among known metabolites and 0.41 (Q1: 0.34; Q3: 0.48) among unknown metabolite features. The most stable (median ICCs: 0.54-0.57) metabolite classes were nucleosides, nucleotides and analogues, phosphatidylcholine plasmalogens, diglycerides, and cholesteryl esters. The least stable (median ICCs: 0.26-0.36) metabolite classes were lysophosphatidylethanolamines, lysophosphatidylcholines and steroid and steroid derivatives. Results in the secondary dataset were similar (Spearman correlation=0.87) to corresponding results in the primary dataset.

**Conclusion:** Within-person stability over 10 years is reasonable for lipid, lipid-related, and polar metabolites, and varies by metabolite class. Additional studies are required to estimate within-person stability over 10 years of other metabolites groups.

## Introduction

In epidemiological studies, samples are often collected long before disease onset or outcome assessment. Within the Nurses’ Health Studies (NHS) and NHSII, several nested case-control studies investigated prospective associations of plasma biomarkers measured in samples collected >10 years before disease onset with risk of developing cancer and other chronic diseases. For example, total circulating carotenoids measured up to 20 years before diagnosis were associated with decreased risk of developing breast cancer^1^. In contrast, high plasma prolactin levels measured ≥10 years before diagnosis were not associated with increased risk of breast cancer while measures <10 years before diagnosis were associated with increased risk^2^, emphasizing that understanding how the timing of biomarkers is related to risk is critical to elucidating disease etiology. Thus, understanding the long-term within-person stability of biomarkers measured in these samples is crucial.

Metabolomics, which reflect the integrated effects of the genetic background, lifestyle and environmental factors^3^, are of particular interest in epidemiologic studies. Metabolites measured up to 23 years before diagnosis were associated with risk of ovarian cancer in NHS and NHSII^4,5^ reflecting long-term disease-biomarker associations. We reported previously on metabolomics short-term within-person stability in these cohorts^6^. However, data on the long-term within-person stability of metabolomics are lacking. Here, we assessed metabolomics within-person stability over 10 years in two separate prospective case-control studies nested within a large epidemiological study, the NHS.

## Methods

### Study Population

In 1976, 121,701 female registered nurses aged 30-55y enrolled in the NHS with the return of a mailed questionnaire^7^. Participants have been followed biennially with questionnaires collecting information on reproductive history, lifestyle factors, diet, medication use, and new disease diagnoses. In 1989-1990, 32,826 NHS participants aged 43-69y contributed blood samples, as previously described^8^. In 2000-2002, 18,473 of these women aged 53-80y donated a second sample using a similar protocol.

The primary dataset was obtained from a prospective breast cancer case-control study nested within the NHS. Incident cases of breast cancer (N=940) were identified after the second blood collection among women who had no reported cancer (other than non-melanoma skin). In total, 1880 women donated a sample during the first blood collection and 1184 women donated a second blood sample approximately 10 years later.

The secondary dataset is from a prospective diabetes case-control study nested within the NHS. Incident cases of diabetes (N=728) were identified after the second blood collection. In total, 1456 women donated samples during the first blood collection and 488 women donated a second blood sample approximately 10 years later.

The study protocol was approved by the institutional review boards of the Brigham and Women’s Hospital and Harvard T.H. Chan School of Public Health, and those of participating registries as required. The return of the self-administered questionnaire and blood sample was considered to imply consent.

### Blood collection methods

The same protocol was used for both blood collections. Briefly, participants had their blood drawn in sodium heparin tubes at a nearby clinic or by their colleagues, and the blood samples were shipped with an ice pack via overnight courier to our laboratory. Whole blood samples were centrifuged (2500 RPM for 20 minutes at 4°C) and aliquoted into 5 mL plasma, red blood cells and white blood cells cryotubes. Plasma samples are stored in the vapor phase of liquid nitrogen (LN2) freezers (temperature ≤ –130°C; alarmed and monitored 24 hours a day) with LN2-rated gasketed screw tops since collection. At the time of blood collection, participants also completed a questionnaire regarding time since last meal and time of day when they completed blood collection

### Laboratory assays

Metabolic profiles were assayed through a metabolomic profiling platform at the Broad Institute using a liquid chromatography tandem mass spectrometry (LC-MS) method designed to measure polar metabolites such as amino acids and lipids^9-11^. The relative abundance of each metabolite was determined by the integration of LC-MS peak areas, which are unitless numbers that are directly proportional to metabolite concentrations. For each measurement method (polar metabolites and lipids), pooled plasma reference samples were included every 20 samples and results were standardized using the ratio of the value of the sample to the value of the nearest pooled reference multiplied by the median of all reference values for the metabolite. In each dataset, samples were assayed in pairs, with matched case-control pairs (as sets) distributed randomly within the batch, and the order of the case and controls within each pair randomly assigned. Therefore, the case and its control were always directly adjacent to each other in the analytic run, thereby limiting variability in platform performance across matched case-control pairs. In addition, YY quality control (QC) samples, to which the laboratory was blinded, were also profiled. These were randomly distributed among the participants’ samples.

Hydrophilic interaction liquid chromatography (HILIC) analyses of water soluble metabolites in the positive ionization mode were conducted using an LC-MS system comprised of a Shimadzu Nexera X2 U-HPLC (Shimadzu Corp.; Marlborough, MA) coupled to a Q Exactive mass spectrometer (Thermo Fisher Scientific; Waltham, MA). Metabolites were extracted from plasma (10 µL) using 90 µL of acetonitrile/methanol/formic acid (74.9:24.9:0.2 v/v/v) containing stable isotope-labeled internal standards (valine-d8, Sigma-Aldrich; St. Louis, MO; and phenylalanine-d8, Cambridge Isotope Laboratories; Andover, MA). The samples were centrifuged (10 min, 9,000 x g, 4°C), and the supernatants were injected directly onto a 150 × 2 mm, 3 µm Atlantis HILIC column (Waters; Milford, MA). The column was eluted isocratically at a flow rate of 250 µL/min with 5% mobile phase A (10 mM ammonium formate and 0.1% formic acid in water) for 0.5 minute followed by a linear gradient to 40% mobile phase B (acetonitrile with 0.1% formic acid) over 10 minutes. MS analyses were carried out using electrospray ionization in the positive ion mode using full scan analysis over 70-800 m/z at 70,000 resolution and 3 Hz data acquisition rate. Other MS settings were: sheath gas 40, sweep gas 2, spray voltage 3.5 kV, capillary temperature 350°C, S-lens RF 40, heater temperature 300°C, microscans 1, automatic gain control target 1e6, and maximum ion time 250 ms. Metabolites measured with this method will be referred to as HILIC-positive metabolites.

Plasma lipids were profiled using a Shimadzu Nexera X2 U-HPLC (Shimadzu Corp.; Marlborough, MA). Lipids were extracted from plasma (10 µL) using 190 µL of isopropanol containing 1,2-didodecanoyl-sn-glycero-3-phosphocholine (Avanti Polar Lipids; Alabaster, AL). After centrifugation, supernatants were injected directly onto a 100 × 2.1 mm, 1.7 µm ACQUITY BEH C8 column (Waters; Milford, MA). The column was eluted isocratically with 80% mobile phase A (95:5:0.1 vol/vol/vol 10mM ammonium acetate/methanol/formic acid) for 1 minute followed by a linear gradient to 80% mobile-phase B (99.9:0.1 vol/vol methanol/formic acid) over 2 minutes, a linear gradient to 100% mobile phase B over 7 minutes, then 3 minutes at 100% mobile-phase B. MS analyses were carried out using electrospray ionization in the positive ion mode using full scan analysis over 200–1100 m/z at 70,000 resolution and 3 Hz data acquisition rate. Other MS settings were: sheath gas 50, in source CID 5 eV, sweep gas 5, spray voltage 3 kV, capillary temperature 300°C, S-lens RF 60, heater temperature 300°C, microscans 1, automatic gain control target 1e6, and maximum ion time 100 ms. Lipid identities were denoted by total acyl carbon number and total double bond number. Metabolites measured with this method will be referred to as C8-positive metabolites.

Raw data from orbitrap mass spectrometers were processed using TraceFinder 3.3 software (Thermo Fisher Scientific; Waltham, MA) and Progenesis QI (Nonlinear Dynamics; Newcastle upon Tyne, UK). For each method, metabolite identities were confirmed using authentic reference standards or reference samples.

After exclusion of metabolites not stable with a delay in processing which is characteristic of the two blood collections^6^ (N=35) and those missing in >10% of the participants who donated two samples (20 known compounds and 1069 unknown metabolite features), the primary dataset included 5938 metabolites (295 known compounds and 5643 unknown metabolite features) measured at both blood collections. 2519 lipids and lipid-related metabolites (153 known compounds and 2366 unknown metabolite features) were measured with the C8 column in positive mode while 3419 polar metabolites (142 known compounds and 3277 unknown metabolite features) were measured with the HILIC column in positive mode. Most of the known metabolites (N=253; 86%) and 47% (N=2655) of the unknown metabolite features had coefficients of variation (CV) <25%. All metabolites were included in this study.

Similarly, after exclusion of metabolites not stable with a delay in processing which is characteristic of the two blood collections^6^ (N=34) and those missing in >10% of the participants who donated two samples (16 known compounds and 427 unknown metabolite features), the secondary dataset included 3209 polar metabolites (202 known compounds and 3007 unknown metabolite features) measured at both blood collections. The secondary dataset did not include lipids and lipid-related metabolites. Most of the known metabolites (N=171; 85%) and 44% (N=1326) of the unknown metabolite features had CV<25%. All metabolites were included in this study.

### Statistical Analysis

Metabolite values were transformed to probit scores within in each blood collection and dataset. We estimated within-person stability over 10 years by calculating intra-class correlation (ICC) using liner mixed models with participant IDs as a random effect. We followed the approach developed by Dr. Rosner et. al^12^ to estimate ICCs on the probit scale and transform these back to the ranked scale. Within-person variability was estimated among participants who donated two blood samples approximately 10 years apart (N=1184) while between-person variability was estimated among all participants (N=1880). ICCs were calculated as the proportion of the total variability (within-person + between-person) attributable to between-person variability. We also calculated median % change in metabolite levels over 10 years. In sensitivity analyses, we restricted to fasting women (N=1309 of which 765 donated 2 samples), women with stable BMI (≤2kg/m^2^ change in BMI between the two blood collections, N=706), women with change in BMI (>2kg/m^2^ change in BMI between the two blood collections, N=478), postmenopausal women not using postmenopausal hormone therapy at either timepoint (N=577 of which 223 donated 2 samples), and to women without breast cancer at both blood collections (N=940 of which 592 donated 2 samples). The secondary dataset included only women who were fasting for >8 hours. To assess differences in ICCs between the two datasets, we calculated % absolute change in ICCs between the primary and secondary dataset.

## Results

### Study population characteristics

Women in the primary dataset had a mean age of 56 years at the first blood collection (N=1880) and 66 years at the second blood collection (N=1184). Participant characteristics were similar at the two blood collections (Table 1) with some exceptions: more women reported being postmenopausal, past smokers and fasting >8 hours at the second blood collection compared to the first blood collection. Women in the secondary dataset (first collection N=1456, second collection N=488) were similar to the fasting women (first collection N=1309, second collection N=1062) in the primary dataset (Supplementary Table 1).

**Table 1:**
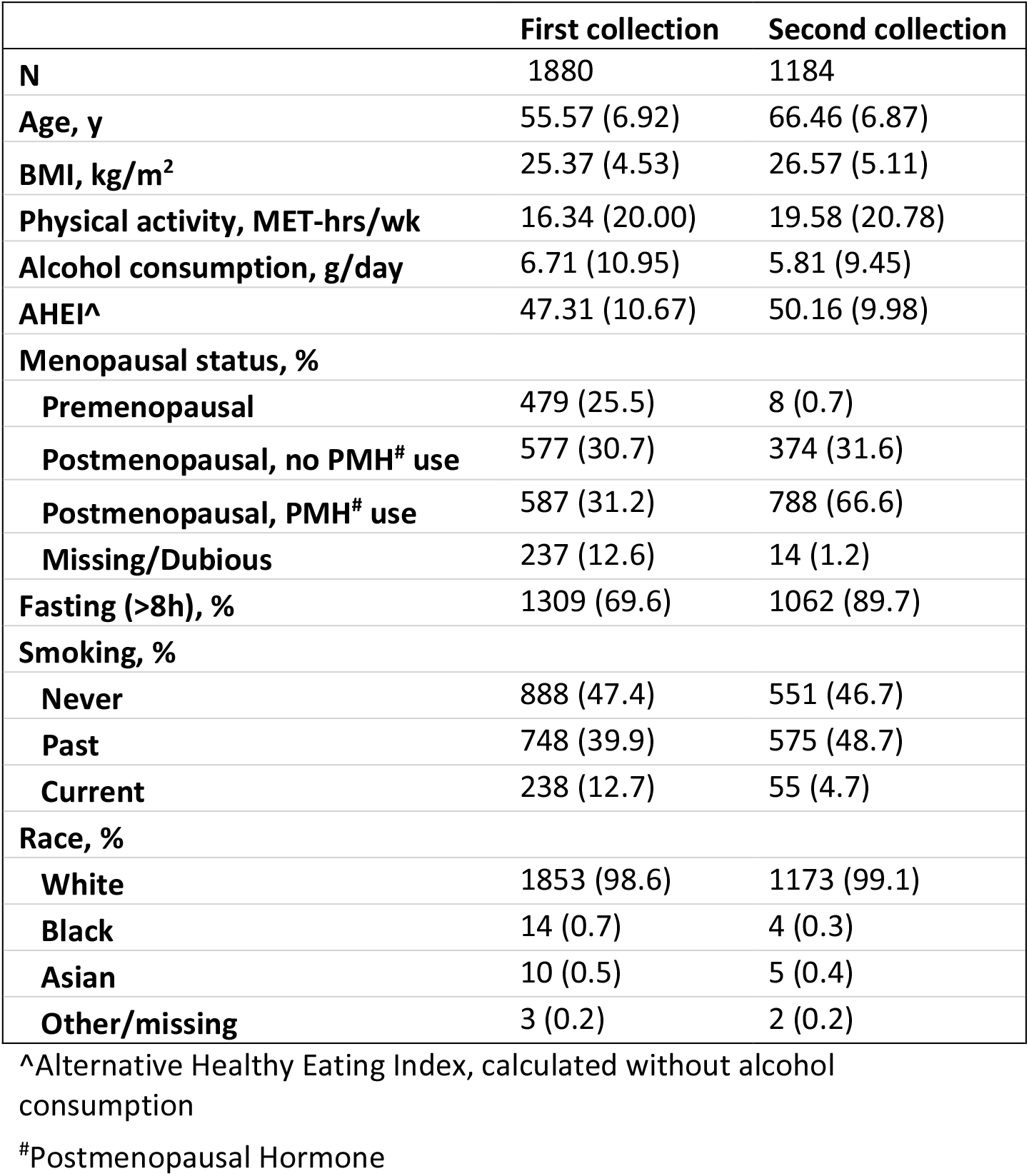
Characteristics of study participants by blood collection in the primary dataset.

### Metabolomics stability over 10 years in the primary dataset

For known metabolites, the median ICC was 0.43 (1^st^ quartile [Q1]: 0.36; 3^rd^ quartile [Q3]: 0.50). For unknown metabolite features, the median ICC was 0.41 (Q1: 0.34; Q3: 0.48). Among lipids and lipid-related metabolites, the median ICC for known metabolites was 0.44 (Q1: 0.38; Q3: 0.51) while for the unknown metabolite features it was 0.41 (Q1: 0.35; Q3: 0.47; Figure 1). Among polar metabolites, the median ICC for known metabolites was 0.42 (Q1: 0.33; Q3: 0.49) while for the unknown metabolite features it was 0.41 (Q1: 0.34; Q3: 0.48; Figure 1). The median ICC was 0.43 (Q1: 0.36; Q3: 0.50) among metabolites with CV<25% and 0.39 (Q1: 0.33; Q3:0.46) among metabolites with CV≥25%. The median % difference in known metabolite levels between the two collections, calculated from the raw values, was -5.25% (Q1: -14.80%, Q3:0.92%; Supplementary Table 2). The median % difference in unknown metabolite feature levels between the two collections, calculated from the raw values, was 0.03% (Q1: -6.80%, Q3:4.13%; Supplementary Table 2).

**Figure 1:**
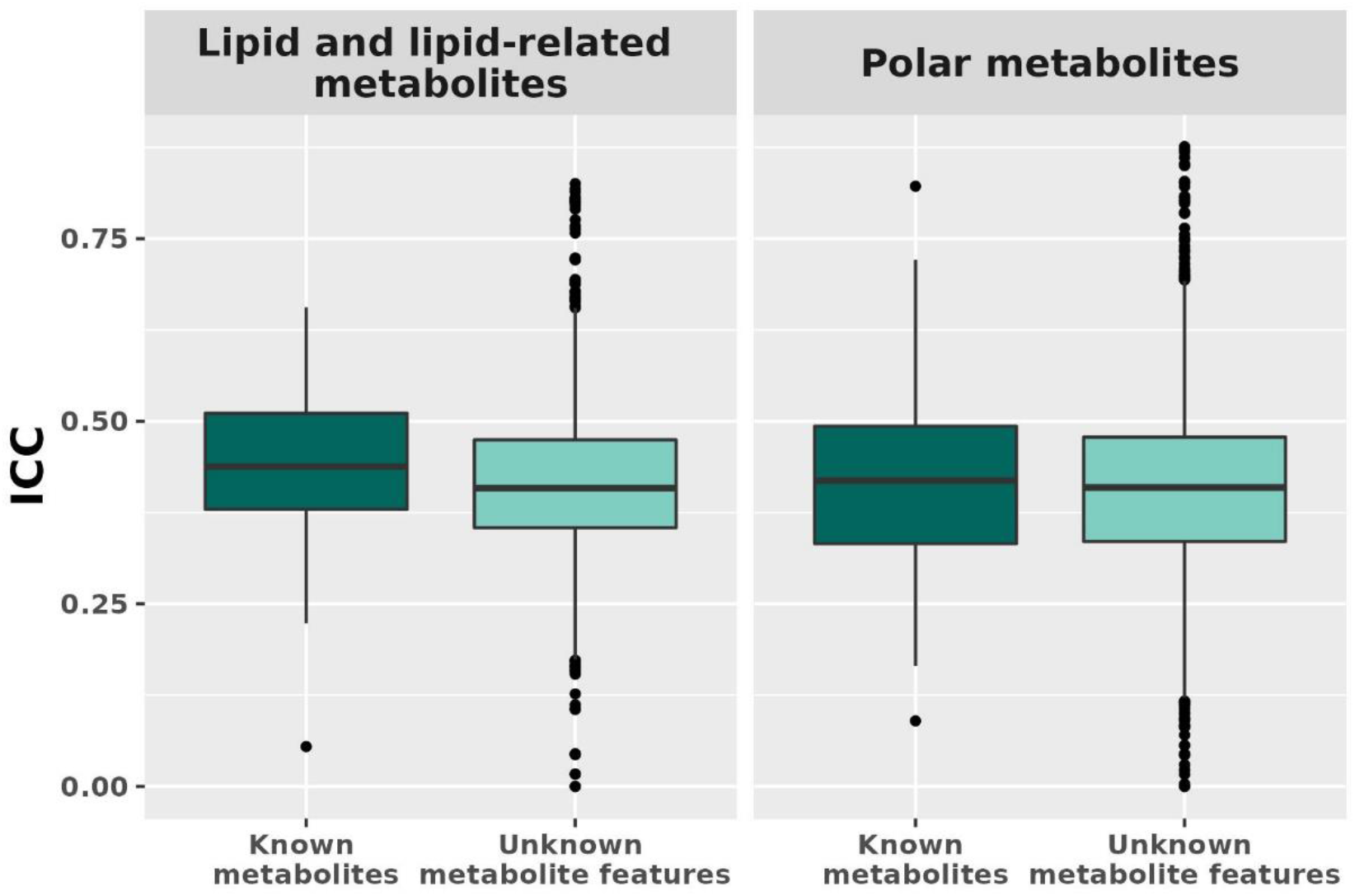
Metabolomics stability over 10 years by measurement method (lipid and lipid-related vs. polar metabolites) and known metabolites (N=295) vs. unknown metabolite features (N=5643).

Among known metabolites, ICCs varied by metabolite class (Figure 2). The most stable metabolite classes were nucleosides, nucleotides, and analogues (median ICC: 0.57; Q1: 0.54; Q3: 0.60), phosphatidylcholine (PC) plasmalogens (median ICC: 0.54; Q1: 0.50; Q3: 0.56), diglycerides (DG; median ICC: 0.53; Q1: 0.51; Q3: 0.54), and cholesteryl esters (CE; median ICC: 0.53; Q1: 0.51; Q3: 0.56). The least stable metabolite classes were steroids and steroid derivatives (median ICC: 0.26; Q1: 0.25; Q3: 0.28), lysophosphatidylcholines (LPC; median ICC: 0.36; Q1: 0.27; Q3: 0.40) and lysophosphatidylethanolamines (LPE; median ICC: 0.36; Q1: 0.33; Q3: 0.38).

**Figure 2:**
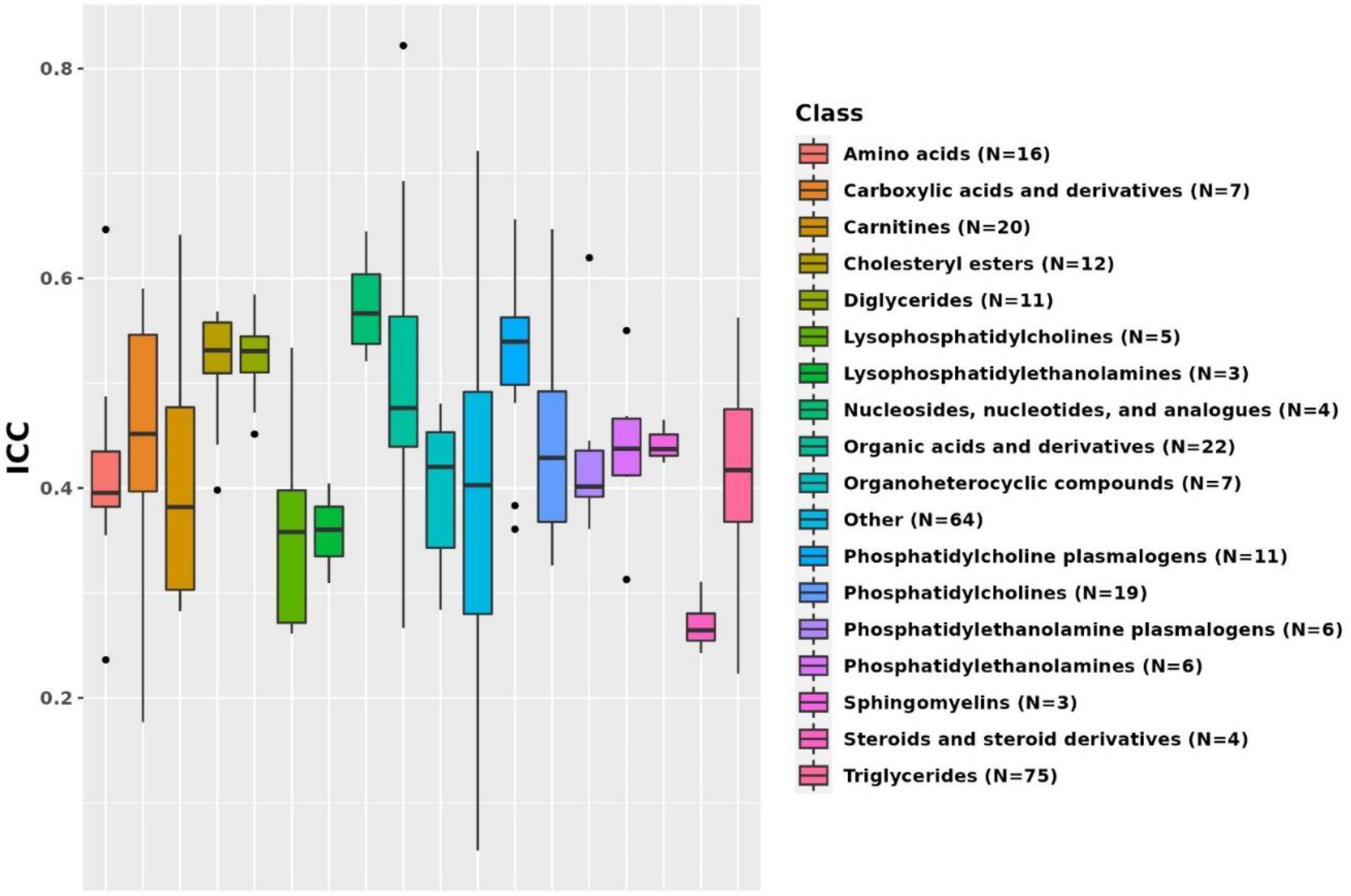
Metabolomics stability over 10 years by metabolite class. Results from known metabolites are included in this figure. Metabolomics classes with less than three metabolites were added to the class Other.

We observed similar ICCs across participant strata in sensitivity analyses (Figure 3, Supplementary Table 2, and Supplementary Figure 1). ICCs of known metabolites estimated among all women (median ICC: 0.43, Q1: 0.36; Q3: 0.50) were similar to those estimated among fasting women (median ICC: 0.45, Q1: 0.37; Q3: 0.52), women with stable BMI (median ICC: 0.43, Q1: 0.36; Q3: 0.51) and women with a change in BMI (median ICC: 0.41, Q1: 0.33; Q3: 0.48). A similar pattern was observed for unknown metabolite features (all women: median ICC: 0.41 (Q1:0.34; Q3:0.48); fasting women: median ICC: 0.42 (Q1: 0.35; Q3: 0.49); women with stable BMI: median ICC: 0.42 (Q1: 0.35; Q3: 0.48); women with unstable BMI: median ICC: 0.40 (Q1: 0.33; Q3: 0.47)). Among postmenopausal women not using postmenopausal hormone therapy at either collection, the median ICC was 0.44 (Q1: 0.36; Q3: 0.53) for known metabolites and 0.41 (Q1: 0.33; Q3: 0.49) for unknown metabolite features (data not shown). Among control samples only, the median ICC was 0.44 (Q1: 0.37; Q3: 0.51) for known metabolites and 0.41 (Q1: 0.35; Q3: 0.49) for unknown metabolite features (data not shown). Notably, metabolites stable among all women were also stable when assessed in different participant strata (for example, N6,N6-dimethyllysine ICC ranges between 0.82 and 0.84 across participant strata; Table 2). Similarly, metabolites with low ICC among all women were also not stable when assessed in the different participant strata (for example, palmitoylethanolamide ICC ranges between 0.03 and 0.06 across subgroups; Table 2).

**Table 2:** Most and least stable metabolites. ICCs were estimated among four participant subgroups: all participants (N=1184 repeated samples and N=696 unique samples), fasting participants (N=765 repeated and N=544 unique samples), and among participants with stable (N=706 repeated samples) or unstable BMI (N=478 repeated samples). The stable BMI group includes participants with ≤2kg/m^2^ difference in BMI between the two blood collections. The unstable BMI group includes participants with >2kg/m^2^ difference in BMI between the two blood collections.

|  |  |  | Intra-class correlation |  |  |  |
| --- | --- | --- | --- | --- | --- | --- |
|  | Metabolite name | Metabolite class | All | Fasting | Stable BMI | Unstable BMI |
| Most stable metabolites | N6,N6-dimethyllysine | Organic acids and derivatives | 0.82 | 0.84 | 0.83 | 0.83 |
|  | DMGV | Other | 0.72 | 0.73 | 0.73 | 0.71 |
|  | N-acetylornithine | Organic acids and derivatives | 0.69 | 0.72 | 0.7 | 0.68 |
|  | C34:2 PC plasmalogen | Phosphatidylcholine plasmalogens | 0.66 | 0.67 | 0.66 | 0.61 |
|  | C38:4 PC | Phosphatidylcholines | 0.65 | 0.66 | 0.66 | 0.63 |
|  | glycine | Amino acids | 0.65 | 0.64 | 0.65 | 0.62 |
|  | C5-DC carnitine | Carnitines | 0.64 | 0.63 | 0.66 | 0.62 |
|  | N4-acetylcytidine | Nucleosides, nucleotides, and analogues | 0.64 | 0.62 | 0.68 | 0.62 |
|  | ADMA/SDMA | Organic acids and derivatives | 0.62 | 0.63 | 0.66 | 0.65 |
|  | C36:1 PE plasmalogen | Phosphatidylethanolamine plasmalogens | 0.62 | 0.62 | 0.64 | 0.59 |
| Least stable metabolites | 1-methylhistidine | Other | 0.21 | 0.18 | 0.21 | 0.25 |
|  | 4-hydroxyhippurate | Other | 0.21 | 0.19 | 0.18 | 0.26 |
|  | acetaminophen | Other | 0.2 | 0.18 | 0.21 | 0.21 |
|  | guanosine | Other | 0.2 | 0.20 | 0.17 | 0.23 |
|  | allantoin | Other | 0.18 | 0.21 | 0.15 | 0.20 |
|  | hydroxyproline | Carboxylic acids and derivatives | 0.18 | 0.12 | 0.17 | 0.17 |
|  | methyl N-methylanthranilate | Other | 0.17 | 0.13 | 0.14 | 0.22 |
|  | trimethylamine-N-oxide | Other | 0.16 | 0.15 | 0.12 | 0.24 |
|  | ectoine | Other | 0.09 | 0.08 | 0.08 | 0.10 |
|  | palmitoylethanolamide | Other | 0.05 | 0.03 | 0.05 | 0.06 |

**Figure 3:**
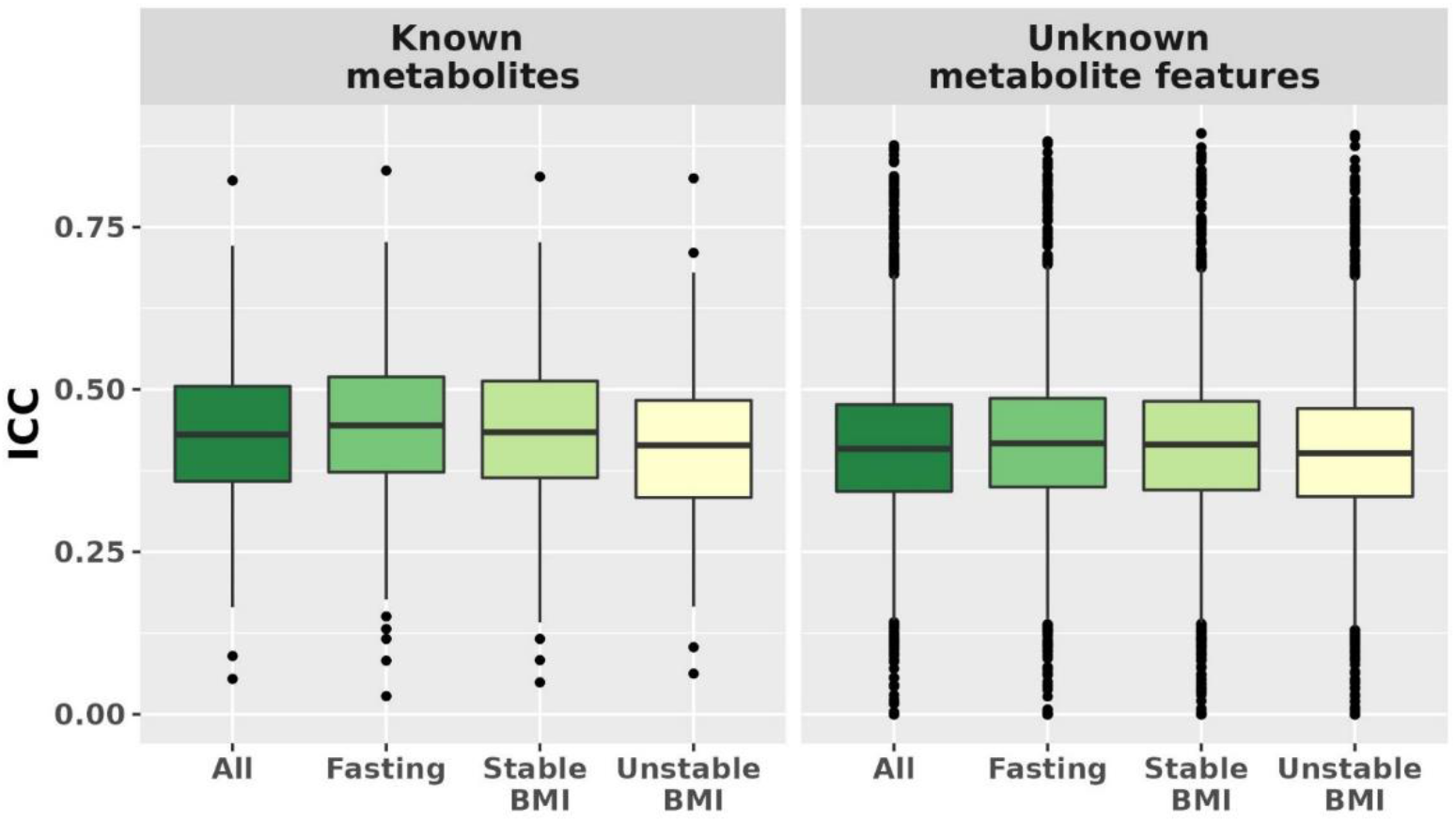
Metabolomics stability over 10 years by known metabolites (N=295) vs. unknown metabolite features (N=5643). ICCs were estimated among all participants (N=1880 of which 1184 donated 2 samples), fasting participants (N=1309 of which 765 donated 2 samples), and among participants with stable (N=706) or unstable BMI (N=478 samples). The stable BMI group includes participants with ≤2kg/m^2^ change in BMI between the two blood collections. The unstable BMI group includes participants with >2kg/m^2^ change in BMI between the two blood collections.

### Metabolomics stability over 10 years in the secondary dataset

Results in the secondary dataset, which included known polar metabolites and fasting women, were similar to results for known polar metabolites among fasting women in the primary dataset (Pearson correlation=0.89, Spearman correlation=0.87; Figure 4, Supplementary Table 3). In the secondary dataset, the median ICC among known metabolites was 0.43 (Q1: 0.34; Q3: 0.51), similar to the median ICC of 0.43 (Q1: 0.35; Q3: 0.52) among unknown metabolite features. Among known metabolites for which we were able to estimate ICCs in both datasets (N=105), 52 (50%) metabolites had % ICC absolute difference <10% and 82 (78%) had % absolute difference <20%. The metabolites with the lowest % absolute ICC difference between the two datasets were 1,7-dimethyluric acid (primary dataset ICC: 0.42; secondary dataset ICC: 0.42), caffeine (primary dataset ICC: 0.42; secondary dataset ICC: 0.43), and creatinine (primary dataset ICC: 0.60; secondary dataset ICC: 0.60). Similarly, 17 metabolites ranked among the top 25 most stable metabolites in the primary dataset were also ranked among the 25 most stable metabolites in the secondary dataset.

**Figure 4:**
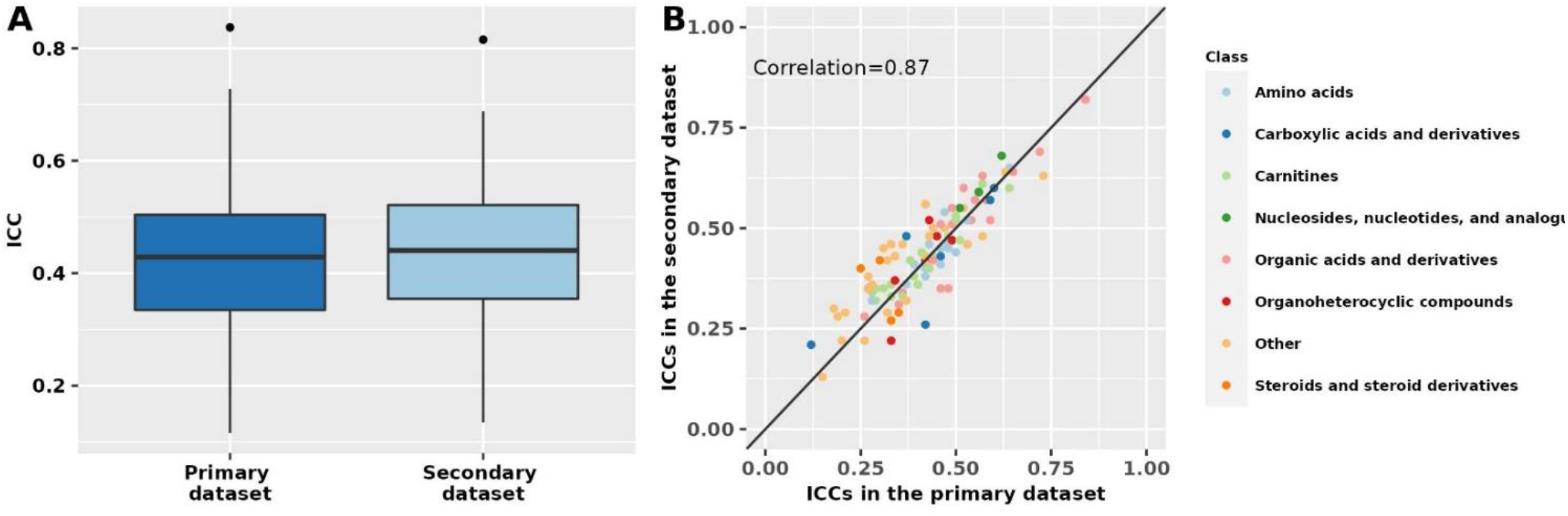
Metabolomics stability over 10 years in the primary and secondary datasets. The primary dataset was restricted to polar metabolites and fasting women to match the secondary dataset. Intra-class correlations (ICCs) in the two datasets are shown as boxplots (panel A) and by metabolite class in a scatter plot (panel B). The correlation was estimated using Spearman’s rank correlation coefficient.

## Discussion

We estimated within-person stability over ten years for 5938 metabolites (295 known compounds and 5643 unknown metabolite features) among 1880 women. Most metabolites were reasonably stable over ten years with a median ICC of 0.43 for known metabolites and 0.40 from unknown metabolite features. Within-person stability over ten years varied by metabolite class. In secondary analyses, we performed a partial replication of polar metabolites in another data set of 1456 fasting women; findings were similar.

To the best of our knowledge, this is the first study to assess within-person stability of metabolomic profiles over 10 years. We have assessed within-person stability of metabolites over shorter periods of times, 1-2 years^6^. Notably, although the within-person stability was attenuated, the metabolite classes that were the most stable over 1-2 years were also the most stable over 10 years. For example, cholesteryl esters, phosphatidylcholine plasmalogens and diglycerides had the high median ICCs over 1-2 years (median ICC: 0.76, 0.76, 0.73) and over 10 years (median ICC: 0.53, 0.55, 0.53). Nucleosides, nucleotides, and analogues were the most stable class over 10y (10y ICC=0.57, 1-2y ICC = 0.55). Steroid and steroid derivatives was the least stable class over 1-2 years (median ICC = 0.36) and over 10 years (median ICC = 0.26).

Our results show that although the within-person stability decreases over time, metabolites are reasonably stable over 10 years. The reduced within-person stability over 10 years compared to 1-2 years is due to multiple sources of variation. True changes in metabolite levels over long periods of time are the most important one. Changes over 10 years in personal, behavioral, and lifestyle factors such as age, BMI, menopausal status, exposure to postmenopausal hormone therapy, diet, physical activity are likely to affect metabolite levels. For example, acetaminophen, a drug often used sporadically for different types of aches and pains, had a low with-in-person stability, likely reflecting different windows of exposure. Furthermore, some diet related metabolites (e.g. trimethylamine-N-oxide (TMO)^13^, pipecolic acid^14^) tended to show low within-person stability over 10 years. Notably, within person stability over time among triglycerides (TAG) varied by saturation level and length of the fatty acyl chains. Most highly unsaturated TAGs with long fatty acyl chains, which are associated with long-term vegetable intake^14^, tended to be more stable over time compared to less unsaturated TAGs with shorter fatty acyl chains. While changes in behaviors and exposures result in a reduced 10 years within-person stability of metabolites, it is important to note that we expect changes in metabolite levels in response to changes in these factors. Furthermore, we study metabolites to identify new risk and disease biomarkers because they reflect changes in these factors and are considered a representation of the metabolic state of an individual, the integrated effects of their genetic background, lifestyle, and environmental exposures^3^. A critical feature of some of the most widely applied clinical disease risk markers, such as standard blood lipids, is their responsiveness to pharmacological and lifestyle-based risk prevention.

Long-term storage may represent another source of variation. However, the samples in these cohorts are stored at ultra-low temperatures (≤-196°C) which were shown to limit the negative effects of long-term storage^15^. Additionally, we have identified metabolites measured in these long-term stored samples that were significantly associated with cancer and other chronic diseases in multiple studies (e.g., pancreatic^16^, ovarian^4,5^, breast^17,18^ and prostate cancer^19^, rheumatoid arthritis^20^ and cardiovascular disease^21^), suggesting that storage time does not substantially impact biomarker-disease associations. Furthermore, for a considerable subset of metabolites (e.g., cotinine, trigonelline, caffeine, pantothenate, C45:3 TAG, C54:9 TAG), we also observed relatively large median % differences over 10 years (−47%/+58%) and, at the same time, reasonably high ICCs (>0.4), suggesting that potential changes in metabolite levels due to long term-storage are similar across individuals.

For a subset of metabolites, two additional sources of variation have to be considered. To leverage the large coverage of the assay, we did not exclude metabolites with low technical reproducibility from the analysis. Furthermore, the samples in this analysis are subject to delayed processing (24-48 hours after sample collection). We excluded metabolites where we have documented variation with a delay in processing^6^, but this information was not available for all known metabolites, and not available for any of the unknown metabolite features. Notably, both factors would result in a potential underestimation of long-term within-person stability.

The 10-year within-person stability of metabolites is similar to other plasma biomarkers measured in the NHS. For example, postmenopausal hormone levels (estradiol ICC = 0.69, testosterone ICC = 0.71, sex hormone-binding globulin ICC = 0.74, and dehydroepiandrosterone sulfate ICC = 0.54)^22^, 25-hydroxyvitamin D (ICC = 0.51)^23^ and prolactin (ICC=0.39)^2^ all showed high or moderate within-person reproducibility over 10 years, whereas dietary biomarkers such as carotenoids (ICCs ranged between 0.3 for β-carotene to 0.54 for lutein and zeaxanthin)^1^ and fluorescent oxidation products (ICCs range from 0.14 to 0.30)^24^ showed moderate or modest long-term within-person stability. While long-term stability should be factored into result interpretation, many of the most predictive and widely used biomarkers have similar within-person stability over 10 years in this cohort. For example, plasma cholesterol has a 10 years ICC of 0.39 and is highly predictive of coronary artery disease risk in our^25-27^ and other cohorts.

Our study has several strengths and limitations. We were able to assess within-person stability over 10 years for over 295 known metabolites (polar metabolites, and lipids and lipid-related metabolites) and 5643 unknown metabolite features. However, our study did not include other metabolite groups such as fatty acids, carbohydrates, alcohols, and vitamins. We had a large sample size and were able to conduct a partial replication for a subset of the measured metabolites among fasting women, but our study included mostly Caucasian women limiting its generalizability. It should also be noted that the samples used in this analysis were subject to delayed processing (blood collection characteristic in the NHS). While we excluded metabolites known to vary with a delay in processing, this information was not available for all analyzed metabolites. Additionally, we did not exclude metabolites with low technical reproducibility. Due to these factors, the ICCs presented here may include variation due to technical reproducibility and/or differential delay in processing between the two blood collections which can potentially result in an underestimation of within-person stability over time.

In summary, our study showed that metabolites are reasonably stable over 10 years, a time interval characteristic of prospective epidemiologic studies of chronic disease. In a secondary dataset, we were able to replicate our findings for a subset of metabolites among fasting women. Stability over ten years varied by metabolite class. While the 10 years stability of metabolites should be an important factor when interpreting results, it is equally important to consider the sources of variation that influence long-term within-person stability of metabolites. Findings from this study represent a comprehensive resource for the design of future studies into disease risk associations of specific metabolites and/or metabolite classes.

## Data Availability

All data produced in the present study are available upon reasonable request to the authors.

## Acknowledgements

This study was funded by the National Institutes of Health (UM1 CA186107, P01 CA087969, R01 CA49449, R01 DK112940). The content is solely the responsibility of the authors and does not necessarily represent the official views of the National Institutes of Health. We would like to thank the participants and staff of the Nurses’ Health Studies for their valuable contributions as well as the following state cancer registries for their help: AL, AZ, AR, CA, CO, CT, DE, FL, GA, ID, IL, IN, IA, KY, LA, ME, MD, MA, MI, NE, NH, NJ, NY, NC, ND, OH, OK, OR, PA, RI, SC, TN, TX, VA, WA, WY. The authors assume full responsibility for analyses and interpretation of these data.

## References

1. Eliassen, A.H., et al. Plasma carotenoids and risk of breast cancer over 20 y of follow-up. Am J Clin Nutr 101, 1197–1205 (2015).

2. Tworoger, S.S., et al. A 20-year prospective study of plasma prolactin as a risk marker of breast cancer development. Cancer Res. 73, 4810–4819 (2013).

3. Krumsiek, J., Bartel, J. & Theis, F.J. Computational approaches for systems metabolomics. Current opinion in biotechnology 39, 198–206 (2016).

4. Zeleznik, O.A., et al. Circulating Lysophosphatidylcholines, Phosphatidylcholines, Ceramides, and Sphingomyelins and Ovarian Cancer Risk: A 23-Year Prospective Study. J Natl Cancer Inst (2019).

5. Zeleznik, O.A., et al. A prospective analysis of circulating plasma metabolites associated with ovarian cancer risk. Cancer Res. (2020).

6. Townsend, M.K., et al. Reproducibility of metabolomic profiles among men and women in 2 large cohort studies. Clinical chemistry 59, 1657–1667 (2013).

7. Hankinson, S.E., et al. Plasma prolactin levels and subsequent risk of breast cancer in postmenopausal women. J. Natl. Cancer Inst. 91, 629–634 (1999).

8. Tworoger, S.S., Sluss, P. & Hankinson, S.E. Association between plasma prolactin concentrations and risk of breast cancer among predominately premenopausal women. Cancer research 66, 2476–2482 (2006).

9. Mascanfroni, I.D., et al. Metabolic control of type 1 regulatory T cell differentiation by AHR and HIF1-α. Nature medicine 21, 638 (2015).

10. O’sullivan, J.F., et al. Dimethylguanidino valeric acid is a marker of liver fat and predicts diabetes. The Journal of clinical investigation 127, 4394–4402 (2017).

11. Paynter, N.P., et al. Metabolic predictors of incident coronary heart disease in women. Circulation 137, 841–853 (2018).

12. Rosner, B. & Glynn, R.J.J.S.i.m. Interval estimation for rank correlation coefficients based on the probit transformation with extension to measurement error correction of correlated ranked data. 26, 633–646 (2007).

13. Bennett, B.J., et al. Trimethylamine-N-oxide, a metabolite associated with atherosclerosis, exhibits complex genetic and dietary regulation. 17, 49–60 (2013).

14. Wang, F., et al. Mapping the Metabolic Profiles of Long-Term Vegetable, Fruit, and Fruit Juice Consumption. 4, 787–787 (2020).

15. Tworoger, S.S. & Hankinson, S.E. Use of biomarkers in epidemiologic studies: minimizing the influence of measurement error in the study design and analysis. Cancer Causes & Control 17, 889–899 (2006).

16. Mayers, J.R., et al. Elevation of circulating branched-chain amino acids is an early event in human pancreatic adenocarcinoma development. Nature medicine 20, 1193–1198 (2014).

17. Zeleznik, O.A., et al. Branched chain amino acids and risk of breast cancer. medRxiv, 2020.2008.2031.20185470 (2020).

18. Zeleznik, O.A., et al. Circulating amino acids and amino acid-related metabolites and risk of breast cancer among predominantly premenopausal women. NPJ breast cancer 7, 1–10 (2021).

19. Dickerman, B.A., et al. A metabolomics analysis of adiposity and advanced prostate cancer risk in the Health Professionals Follow-up Study. Metabolites 10, 99 (2020).

20. Chu, S.H., et al. Circulating plasma metabolites and risk of rheumatoid arthritis in the Nurses’ Health Study. Rheumatology 59, 3369–3379 (2020).

21. Li, J., et al. The Mediterranean diet, plasma metabolome, and cardiovascular disease risk. European heart journal 41, 2645–2656 (2020).

22. Zhang, X., Tworoger, S.S., Eliassen, A.H. & Hankinson, S.E. Postmenopausal plasma sex hormone levels and breast cancer risk over 20 years of follow-up. Breast cancer research and treatment 137, 883–892 (2013).

23. Eliassen, A.H., et al. Plasma 25-hydroxyvitamin D and risk of breast cancer in women followed over 20 years. Cancer Res. 76, 5423–5430 (2016).

24. Fortner, R.T., Tworoger, S.S., Wu, T. & Eliassen, A.H. Plasma florescent oxidation products and breast cancer risk: repeated measures in the Nurses’ Health Study. Breast cancer research and treatment 141, 307–316 (2013).

25. Cahill, L.E., Sacks, F.M., Rimm, E.B. & Jensen, M.K. Cholesterol efflux capacity, HDL cholesterol, and risk of coronary heart disease: a nested case-control study in men. J. Lipid Res. 60, 1457– 1464 (2019).

26. Jensen, M.K., Rimm, E.B., Furtado, J.D. & Sacks, F.M. Apolipoprotein C-III as a potential modulator of the association between HDL-cholesterol and incident coronary heart disease. Journal of the American Heart Association 1, e000232 (2012).

27. Mendivil, C.O., Rimm, E.B., Furtado, J., Chiuve, S.E. & Sacks, F.M. Low-density lipoproteins containing apolipoprotein C-III and the risk of coronary heart disease. Circulation 124, 2065– 2072 (2011).

